# Deep learning based models to study the effect of glaucoma genes on angle dysgenesis in-vivo

**DOI:** 10.1101/2021.12.18.21267894

**Authors:** Viney Gupta, Shweta Birla, Toshit Varshney, Bindu I Somarajan, Shikha Gupta, Mrinalini Gupta, Karthikeyan Mahalingam, Abhishek Singh, Dinesh Gupta

## Abstract

**Objective:** To predict the presence of Angle Dysgenesis on Anterior Segment Optical Coherence Tomography (ADoA) using deep learning and to correlate ADoA with mutations in known glaucoma genes.

**Design:** A cross-sectional observational study.

**Participants:** Eight hundred, high definition anterior segment optical coherence tomography (ASOCT) B-scans were included, out of which 340 images (One scan per eye) were used to build the machine learning (ML) model and the rest were used for validation of ADoA. Out of 340 images, 170 scans included PCG (n=27), JOAG (n=86) and POAG (n=57) eyes and the rest were controls. The genetic validation dataset consisted of another 393 images of patients with known mutations compared with 320 images of healthy controls

**Methods:** ADoA was defined as the absence of Schlemm’s canal(SC), the presence of extensive hyper-reflectivity over the region of trabecular meshwork or a hyper-reflective membrane (HM) over the region of the trabecular meshwork. Deep learning was used to classify a given ASOCT image as either having angle dysgenesis or not. ADoA was then specifically looked for, on ASOCT images of patients with mutations in the known genes for glaucoma (*MYOC, CYP1B1, FOXC1* and *LTBP2*).

**Main Outcome measures:** Using Deep learning to identify ADoA in patients with known gene mutations.

**Results:** Our three optimized deep learning models showed an accuracy > 95%, specificity >97% and sensitivity >96% in detecting angle dysgenesis on ASOCT in the internal test dataset. The area under receiver operating characteristic (AUROC) curve, based on the external validation cohort were 0.91 (95% CI, 0.88 to 0.95), 0.80 (95% CI, 0.75 to 0.86) and 0.86 (95% CI, 0.80 to 0.91) for the three models. Amongst the patients with known gene mutations, ADoA was observed among all the patients with *MYOC* mutations, as it was also observed among those with *CYP1B1, FOXC1* and with *LTBP2* mutations compared to only 5% of those healthy controls (with no glaucoma mutations).

**Conclusions:** Three deep learning models were developed for a consensus-based outcome to objectively identify ADoA among glaucoma patients. All patients with *MYOC* mutations had ADoA as predicted by the models.

## Introduction

Anterior segment Spectral domain–optical coherence tomography (SD-OCT) is being increasingly used in glaucoma patients, primarily to investigate the anterior chamber angle and visualize the trabecular meshwork (TM) and Schlemm’s canal (SC) in vivo.^1-4^ This in vivo imaging of anterior chamber angle with ASOCT has been used to detect gross features of angle dysgenesis in primary congenital glaucoma (PCG), juvenile onset open angle glaucoma (JOAG) and adult onset primary open angle glaucoma (POAG), which has been described either as an absence of SC and/or the presence of abnormal tissue or a hyper-reflective membrane within angle recess.^5-8^ These studies have shown that angle dysgenesis on ASOCT (ADoA), can be observed even in eyes with gonioscopically normal appearing angles. Primary congenital glaucoma, JOAG and adult onset POAG form a spectrum in terms of severity of angle dysgenesis. While most of the PCG eyes have features of ADoA, the same are present only in 40% of JOAG eyes and in up to 35% of adult onset POAG.^6,8^ Since there exists a wide spectrum of anatomical variability of the drainage angle, the TM and SC morphology in normal eyes^6-8^ which can make it difficult to distinguish normal from abnormal, therefore, interpretation of ASOCT images requires expertise and a deep understanding of the complexity involved in the developmental anomalies of the outflow pathways. But the number of human experts to infer images and refer patients for specialised care is limited and does not match the extensive number of ASOCT imaging now being routinely done.

Artificial intelligence (AI) has the potential to assist experts in disease diagnosis, progression and management by performing rapid image classification, which otherwise is a difficult or ambiguous scenario for human experts. Deep learning (DL), a subtype of AI, uses the concept of biological neural networks and has demonstrated convincing results in ophthalmic diseases.^9-11^

While angle dysgenesis is associated with developmental immaturity of the outflow pathways regulated by genes, this could only be ascertained with the help of histopathological studies. Mutations in some of the commonly associated genes with glaucoma, namely *CYP1B1*^*12*^, *FOXC1*^*13*^, PITX2^14^, and TEK^15^ have been shown to be associated with developmental abnormalities in the outflow pathways in experimental studies. The severity of angle dysgenesis has been correlated on histopathology with certain *CYP1B1* gene mutations in PCG patients.^16^ Though *MYOC* mutations are known to be associated with early onset glaucoma of PCG^17-19^ and JOAG^20-24^ no studies have shown the involvement of *MYOC* mutations in causing angle dysgenesis.Histopathological studies for angle dysgenesis in human glaucomatous eyes are difficult to perform and are inherently associated with tissue handling artefacts. While grossly identifiable features of ADoA have been described before,^6,8^ many subtle changes may also be present on ASOCT scans which are challenging to detect or precisely quantify by human observers. This study was undertaken to identify angle dysgenesis with the help of AI and use it to predict the presence and absence of ADoA among patients with known gene mutations.

## Material and methods

### Dataset details and study design

The study adhered to the tenets of the declaration of Helsinki and was approved by the Institutional Ethics Committee. An informed consent to participate was taken from all cases and healthy subjects. A detailed history was recorded and all subjects underwent a thorough clinical examination.

### Inclusion criteria

#### Normal eyes

Healthy subjects (age > 10years), who had IOP in the normal range, gonioscopically normal open angle and no other ocular pathology on detailed ophthalmic evaluation.

#### PCG

These were unrelated cases of PCG with enlarged corneal diameters (>12 mm) who had baseline IOP records of >22 mm Hg detected before 3 years of age and were now old enough (>10 years of age) to cooperate for anterior segment OCT scanning.

#### JOAG

These were unrelated primary open angle glaucoma patients diagnosed between 10 and 40 years of age.

#### Adult-Onset POAG

These were unrelated cases of POAG diagnosed after the age of 40 years with untreated IOP >22 mm Hg in one or both the eyes on more than two occasions, open angle on gonioscopy in both eyes, and glaucomatous optic neuropathy in one or both eyes with visual field loss consistent with optic nerve damage.

Only those patients who had been treated and had an IOP<22 mmHg at the time of imaging were included.

### Exclusion criteria

Patients excluded from the study: those with a history of steroid use, presence of any other retinal or neurologic pathology, evidence of secondary causes of raised IOP such as pigment dispersion, pseudoexfoliation, or trauma, those with any pathology detected on gonioscopy such as angle recession, pigmentation of the angle greater than grade 3, irido trabecular contact or peripheral anterior synechiae, and patients with nystagmus/and or poor fixation were excluded.

### SD-OCT examination

The OCT examination was performed using the Spectralis OCT (software version 6.5; Heidelberg Engineering GmbH, Heidelberg, Germany). This machine uses an 880-nm wavelength and provides a resolution of 3.5 μm (digital) to 7 μm (optical) at 40 kHz. An anterior segment lens was used. Only those images that were considered good quality were included. ASOCT B-scans from nasal/temporal quadrant were selected per eye and these images were analyzed by 2 blinded observers for presence or absence of ADoA, which were then subsequently used for machine learning. A total of 800 ASOCT B-scans were included, out of which 340 images (1 B-scan per eye) were used to build the ML model and the rest were used for validation. Out of 340 images, 170 scans included PCG (n=27), JOAG (n=86) and POAG (n=57) eyes and the rest were healthy controls.

### Data preprocessing

**Figure 1** summarizes the workflow used in the study. The images were encoded by removing patient details and giving unique reference numbers. Each image was cropped manually in two ways to extract the iridiocorneal angle (ICA) area and a trabecular meshwork (TM) area by a single observer. For this study, the ICA area was defined as 1100 × 900 pixels ± 10% (1210 × 990µm) rectangular area including the region covering TM, SC, a part of cornea in continuation with a part of sclera and iris. The images were further cropped to get a TM area defined as 600 × 400 pixels ± 10% (660 × 440 µm), including SC, scleral spur, and TM region (**Figure 2**).

**Fig 1:**
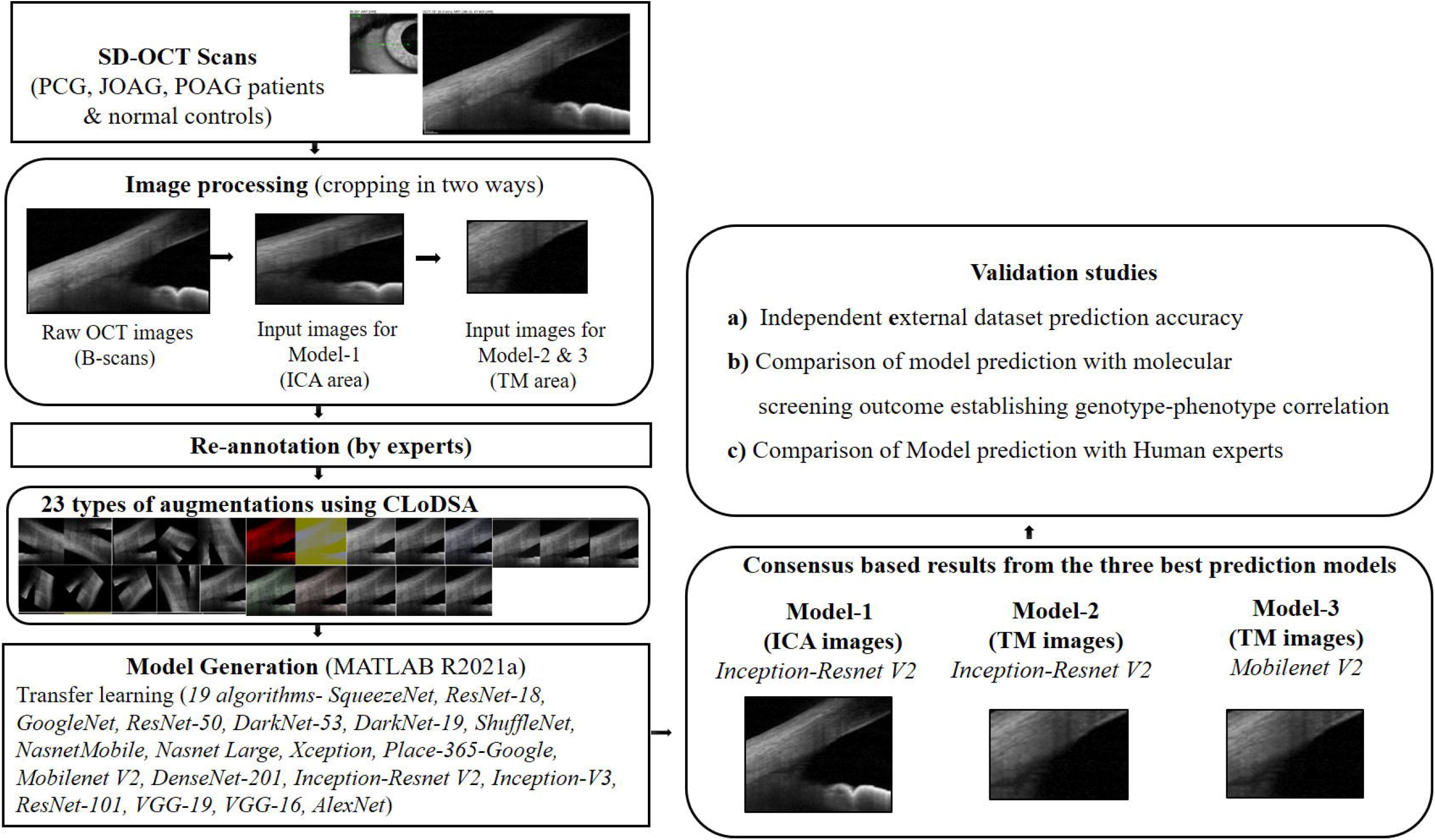
Work flow used in deep learning of anterior segment SD OCT images.

**Fig 2:**
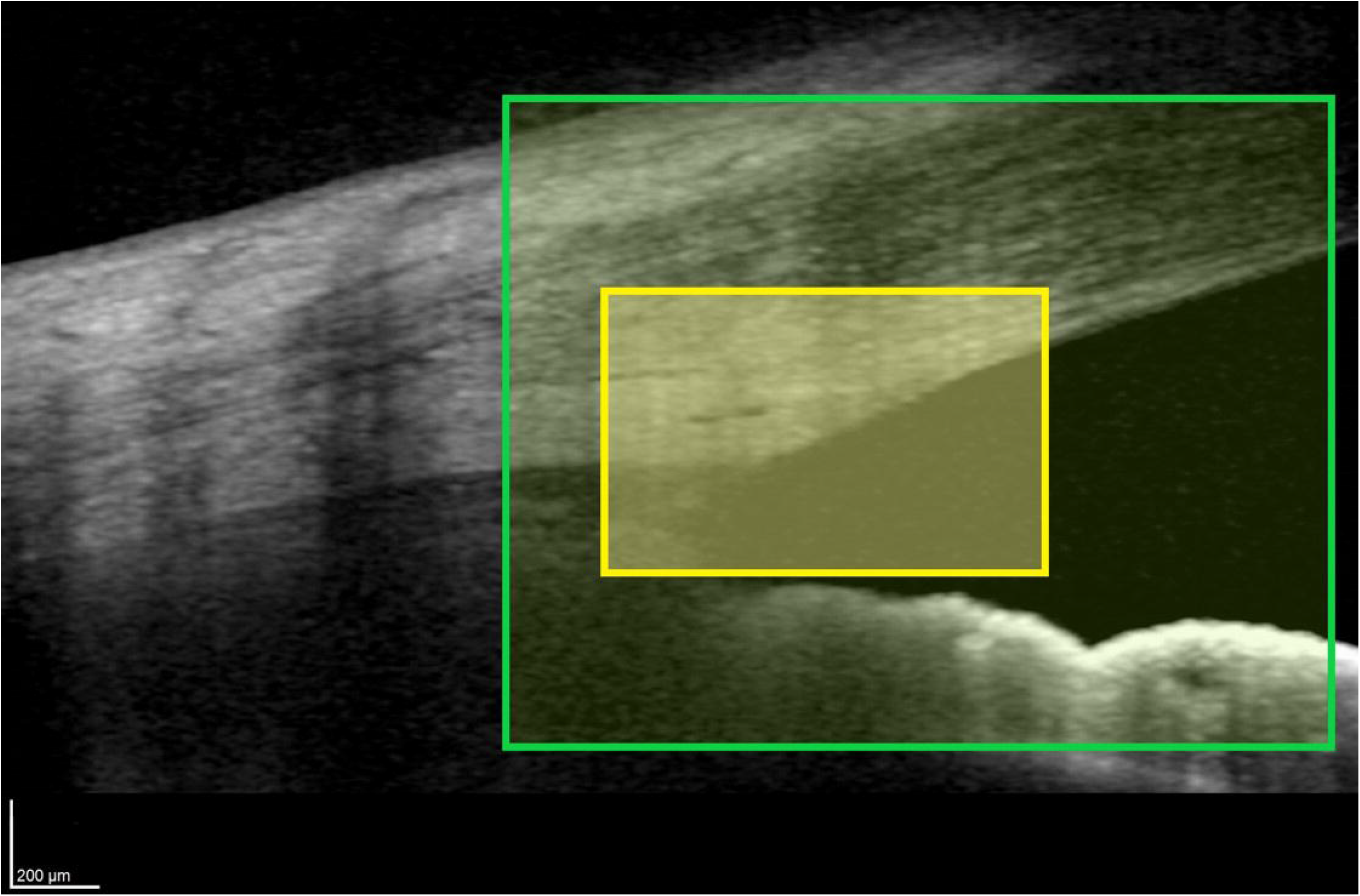
Anterior segment SD OCT image showing the iridocorneal angle area (green rectangle) and the trabecular meshwork area (yellow rectangle).

### Model training and evaluation

#### Augmentation technique and the technology used are provided in Supplemental information 1

The two datasets (ICA area and TM area images), each having 8160 images, were randomly split into training (n=7996) and testing (n=164) subsets with a ratio of 98:2 (**Supplemental Fig 1**). The applied split ratio was considered so that the maximum number of images could be used for model training. The test set was used only for the final evaluation of the model performance and none of the images in the test set were used for training.

We applied the transfer learning method to classify a given SD-OCT image as either having a normal angle or angle dysgenesis. In MATLAB all the available 19 pre-trained convolutional neural network (CNN) models including SqueezeNet, ResNet-18, GoogleNet, ResNet-50, DarkNet-53, DarkNet-19, ShuffleNet, NasnetMobile, Nasnet Large, Xception, Place-365-Google, Mobilenet V2, DenseNet-201, Inception-Resnet V2, Inception-V3, ResNet-101, VGG-19, VGG-16 and AlexNet were trained using our datasets. The first input layer and last output layer with the soft-max activation function in the models were replaced for the binary classification between angle dysgenesis and normal angle. All the images were resized to the required pixels depending upon the CNN model being trained. Initially, all the models were trained using the default parameters and the most efficient ones were prioritised. The hyperparameters of the prioritised models were further tuned in a stochastic gradient descent manner (SGDM) based on minimization of mean squared error with the combinations of different batch sizes, epochs, learning rates, momentum and drop factor.

To get the robust models, different groupings of 23 augmented images were also evaluated along with models with single augmented images and models with all combined augmented images. Finally, 74 models were developed using varied hyper-parameters and augmented images combinations. Prediction quality was assessed by overall accuracy, specificity, sensitivity, area under the ROC curve and comparison with the image annotations of the two experts.

### Genetic correlation with ADoA

Thirty unrelated patients with open angle glaucoma diagnosed between 10 to 40 years of age, who had undergone Whole Exome Sequencing (WES) followed by a bioinformatics analysis (*provided in Supplemental information)* and had been found to harbour a mutation (that was pathogenic) in a known glaucoma gene, underwent ASOCT. The DL models were applied to detect ADoA.

### Validation of DL predictions

Validations of the final DL models were performed using the following types of independent datasets (**Supplemental Fig 1**). a) An independent external validation dataset of 67 images, b) A comparative validation between the model prediction and human experts where two glaucoma specialists (with more than 20 years’ experience) masked to details of the patients, evaluated the SD-OCT scans (n=73) and their results compared with the final model prediction and c) A genetics validation dataset consisted of 393 images of patients with known mutations and 320 images of healthy controls without any glaucoma gene mutation. This was a blind check validation where the results of the molecular analysis were blinded from the AI experts.

### Statistical analysis

To compare the outcomes from the validation studies, statistical analyses were performed using a statistical software package (SPSS v. 26.0; SPSS, Inc., Chicago, IL, USA). To determine the agreement between the specialists and DL prediction, Cohen’s κ test was applied. The Receiver Operating Characteristic (ROC) analysis was performed for the external validation dataset and area under ROC (AUROC) curve was determined for comparisons.

## Results

**Table1** shows the clinical and demographic characteristics of the study subjects.

**Table1:** Demographic and clinical details of subjects whose ASOCT B-scans (1 B-scan image per eye, n = 340) were used for machine learning model preparation.

| Characteristics | PCG | JOAG | POAG | Normal (Control) |
| --- | --- | --- | --- | --- |
| Number of subjects | 16 | 62 | 37 | 85 |
| Number of eyes | 27 | 86 | 57 | 170 |
| Laterality |  |  |  |  |
| • Bilateral | 11 | 24 | 20 | 85 |
| • Unilateral | 5 | 38 | 17 | 0 |
| Gender |  |  |  |  |
| • Male | 5 (31.25%) | 44 (71%) | 25 (67.5%) | 45 (53%) |
| • Female | 11 (68.75%) | 18 (29%) | 12 (32.5%) | 40 (47%) |
| IOP mmHg at the time of study (Mean ± SD) | 14.2 ± 1.4 | 13.8 ± 1.2 | 15.1 ± 1.3 | 16.2 ± 0.8 |

ASOCT – Anterior Segment Optical Coherence tomography, IOP – Intraocular pressure

Using transfer learning on two approaches as mentioned below, we built a consensus-based algorithm consisting of the three best models for differentiating angle dysgenesis from the normal angle. The performance measures of these models are given in **Supplemental Table1**.

The first approach uses the iridocorneal angle area dataset to train all the 19 CNN models (**Figure 1**). The most efficient model was built using Inception-ResnetV2, a 164 layers deep convolutional neural network previously trained on more than a million images.^25^ All the images were rescaled to 299×299 pixels as the input image pre-requisite of Inception-ResnetV2 and finally utilizes SGDM optimiser with a learning rate of 0.005, 45 epochs, and mini-batch size of 64 after hyper parameters optimisation (**Supplemental Table1**). The model achieved the accuracy, sensitivity and specificity on the internal test dataset of 97.56%, 96.4% and 98.7%, respectively.

The second approach uses the TM area and the two best models were trained using Inception-ResnetV2^25^ and Mobilenetv2^26^ neural networks. The mobilenetv2 is a convolutional neural network that requires an input image of 229×229 pixels. Using the TM area test images, the models achieved an accuracy of 98.17% and 98.78%, with a sensitivity of 97% and 98.7%, a specificity of 98.7% in each of the cases, respectively (**Supplemental Table1**). The consensus based outcome from the three CNN transfer learning models is the final predicted classification which could recognise pixel patterns corresponding to the abnormalities at the angle, helping in better classification among the glaucoma group and controls.

### External Validation dataset

To further evaluate the accuracy and reproducibility of our models, we tested them on an independent external validation dataset consisting of 67 images. The models trained with the combined augmented and the original images exhibited lower accuracy than those trained on original images alone. So we did not proceed with the augmented images and all the validation studies were carried using original images only. Model 1 had the best accuracy and specificity but the lowest sensitivity, while the other two models showed good sensitivity and comparable accuracies (**Supplemental Table1**). The consensus-based outcome ensures inclusiveness of the mandatory training features after trade-off and reaching one outcome. The area under the ROC curves were >0.80 for all the three models indicating good performances of the models in detecting ADoA (**Figure 3**).

**Fig 3:**
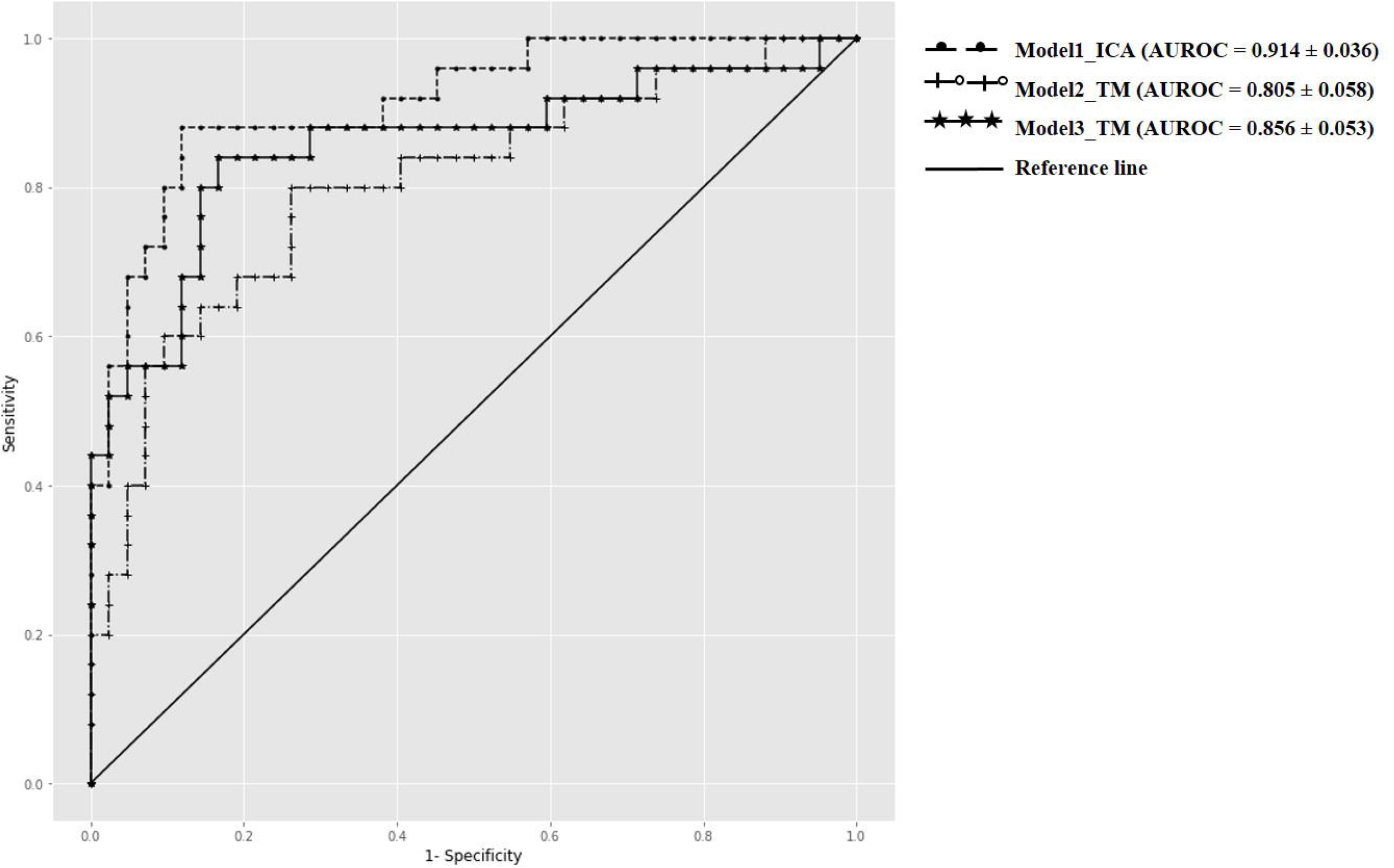
Receiver operating characteristic (ROC) curves for the three Deep Learning models using external validation dataset. *AUROC= Area under ROC curve, TM =Trabecular meshwork, ICA = Iridocorneal angle

### Comparison of the Models performance with Human Experts

The comparative prediction analysis is summarised in **Table 2**. The consensus-based result achieved a maximum accuracy of 83%, reiterating the importance of consensus-based decision-making in clinical settings. To determine the agreement between the expert’s decision and consensus-based prediction, Cohen’s Kappa test was carried out between expert1-model prediction, expert2-model prediction and expert1-expert2 prediction. There was a good agreement between the expert1-model prediction (κ = .619, p<0.05), which is indicative of a similarity between the well experienced expert’s decision and consensus-based model prediction (**Table 3**).

**Table 2:** Comparison between the model prediction and expert’s decision.

|  | **M1** | **M2** | **M3** | **Final consensus-based prediction** |
| --- | --- | --- | --- | --- |
| **Expert1** | 80.82% | 79.45% | 80.82% | 83.56% |
| **Expert2** | 67.12% | 79.45% | 78% | 72.6% |

**Table 3:** Showing Cohen’s κ test results determining the agreement between the experts and model prediction (p < 0.05 was considered significant)

|  | Value | Asymptomatic<br>standard error<sup>a</sup> | Approximate<br>significance |
| --- | --- | --- | --- |
| Expert1/Consensus-based prediction |  |  |  |
| Measurement of<br>agreement Kappa | 0.619 | 0.099 | 0.00 |
| Number of valid cases | 73 |  |  |
| Expert2/Consensus-based prediction |  |  |  |
| Measurement of<br>agreement Kappa | 0.230 | 0.117 | 0.027 |
| Number of valid cases | 73 |  |  |
| Expert1/Expert2 |  |  |  |
| Measurement of<br>agreement Kappa | 0.417 | 0.109 |  |
| Number of valid cases | 73 |  | 0.000 |

a. Not assuming the null hypothesis.

### Genetic validation dataset

Out of 30 (unrelated) patients, who had known gene mutations, 16 had *MYOC* mutations, 10 had *CYP1B1*, 2 had *FOXC1* and 2 had *LTBP2* mutation. The detailed genotype of these patients is provided in **Table 4**. These 30 patients had 16 different mutations, all except three (that were frameshift) were missense. All mutations except one in *CYP1B1* gene (p.Arg368His) were heterozygous.

**Table 4:**
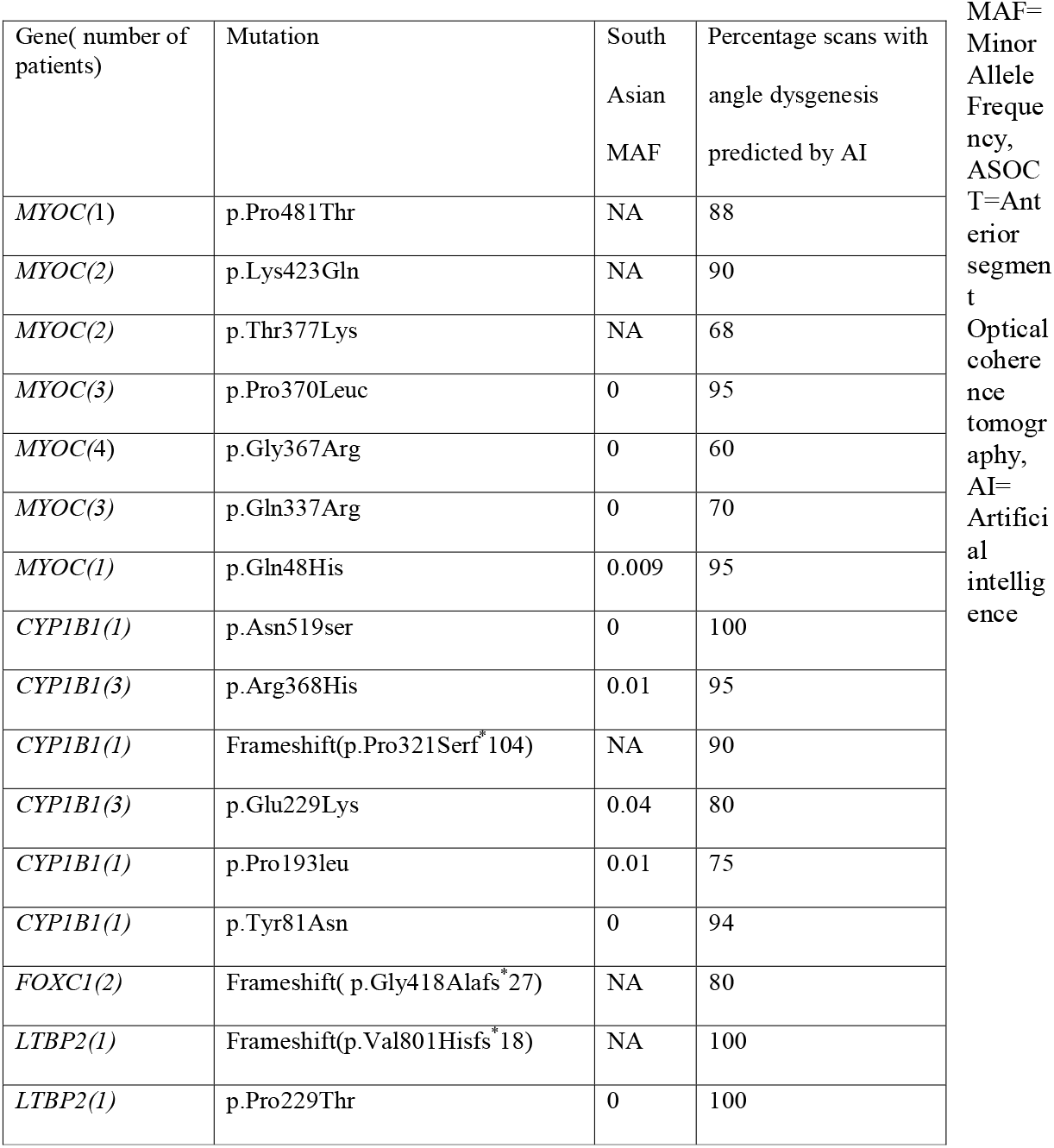
Genetic mutations among 30 patients and angle dysgenesis on ASOCT as determined by AI consensus.

Among these patients, angle dysgenesis on ASOCT was observed as determined by AI, among all patients with known gene mutations. Maximum number of scans showing ADoA were observed with *MYOC* p.Pro370Leuc and p.Gln48His, with *CYP1B1* p.Asn519ser and p.Arg368His and with *LTBP2* frameshift (p.Val801Hisfs*18) and p.Pro229Thr mutation. Gonioscopically angle dysgenesis was not seen among any of the *MYOC* patients. However, features of angle dysgenesis were seen both on gonioscopy and on ASOCT among those with *CYP1B1, FOXC1* and *LTBP2* mutation.

Overall, AI was predictive of angle dysgenesis in 81% scans among *MYOC* positive patients, 89% *CYP1B1* patients, 85% *FOXC1* and 96% among those with *LTBP2* mutation on an average.

**Figure 4** shows images of patients with known gene mutations predicted to have angle dysgenesis on AI Modelling. While *CYP1B1* and *LTBP2* were seen to primarily affect SC morphology, the *MYOC* and *FOXC1* mutations were found to be associated with morphological variations in the TM, since the SC was visualised on most scans. The age of onset of glaucoma did not correlate with the extent of angle dysgenesis.

**Fig 4:**
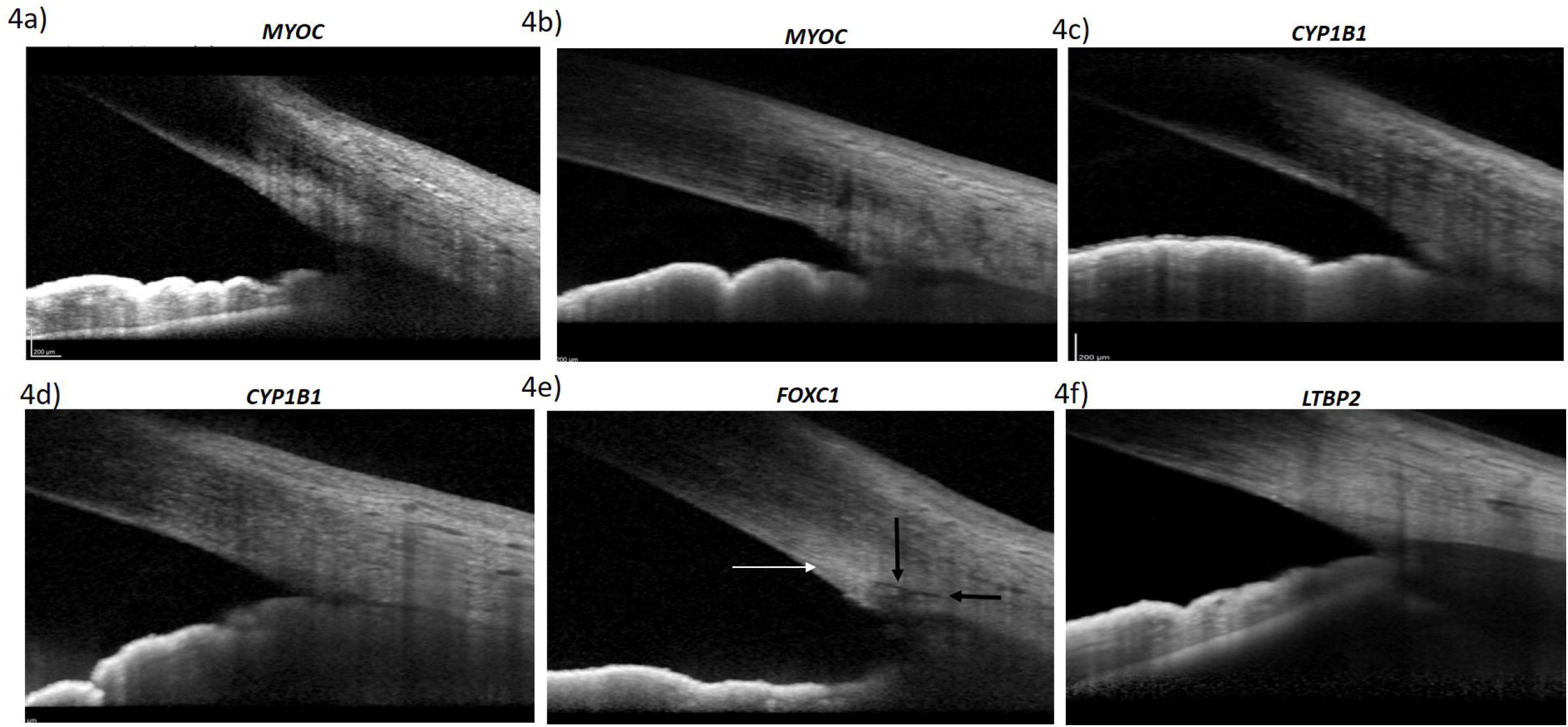
Anterior segment SD OCT images of patients with a)*MYOC* p.Gly367Arg showing intense hyper reflectivity at the TM, b) *MYOC* p.Gln48His showing intense hyper reflectivity at the TM with absent SC, c) *CYP1B1* p.Asn519ser showing intense hyper reflectivity at the TM with absent SC, d) *CYP1B1* p.Tyr81Asn showing absent SC, e) *FOXC1* p.Gly418Alafs*27 showing intense hyper reflectivity at the TM(White arrow) with presence of SC (Black arrows), f) *LTBP2* p.Pro229Thr showing intense hyper reflectivity at the TM with absent SC.

## Discussion

This study used deep learning to build models that identify ADoA among open-angle glaucoma patients. We hypothesised that the angle dysgenesis, exhibited as disturbances in the extracellular matrix of TM, SC and adjoining regions in open angle glaucoma patients, could be reflected as pixel changes (as they occur on histopathological sections) and the DL based models can identify these pixel variations classifying the iridocorneal angle as having dysgenesis or not, based on SD-OCT scans. In the present study, SD-OCT scans of normal, PCG, JOAG and POAG eyes were used to develop a robust deep learning based model by using an image-to-classification approach. In a subset of these patients, ADoA was correlated with known gene mutations.

High definition ASOCT can pick up anomalies in SC development and TM morphology, which are not visible on enface gonioscopy. Gonioscopy and goniophotography also require much expertise and remain observer-dependent. Also sometimes what appears to be a normal open angle may be harbouring dysgenesis in the form of an impermeable hyper reflective membrane that may be visible only on an ASOCT.^8^ While ASOCT has been used for identifying angle closure, it would be of use even in patients with POAG in identifying ADoA, and thus to decide the role of angle based surgeries.^8^

In the present study, three deep learning based models were developed for a consensus based outcome to predict the presence of ADoA among open angle glaucoma patients. Out of all the 19 transfer learning algorithms used in model building, inception resnetv2 and mobilenetv2 achieved superior performances on the ICA area and TM. Whether an eye has angle dysgenesis or not was predicted with greater than 95% accuracy in the internal dataset and more than 80% accuracy in the two different external validation datasets used in the present study.

Considering the phenotypic, genotypic and histopathological complexity in open angle glaucoma, DL has been implemented using different approaches.^27-30,30,31^ Studies have evaluated the potential of implementing DL in primary angle closure disease and have shown promising results.^32,33^ While the SD-OCT scans can successfully capture the anterior angle at high resolution, they may fail to identify angle dysgenesis in cases where altered extracellular matrix anomalies are subtle enough to get unnoticed by human eyes. While identifying gross dysgenesis of the angle, as in PCG may be easier^34^, subtle angle anomalies as in JOAG or POAG are more challenging to identify. The biological changes in the extracellular matrix (ECM) comprising of the trabecular drainage pathways that lead to IOP elevation need to be identified in vivo.^35,36^ With the DL models used in the present study, we could pick up these subtle ECM changes in the TM along with abnormalities in the SC morphology.

Deep learning requires an enormous number of annotated training data, which is challenging to obtain in rare disorders^37^, however, transfer learning and augmentation techniques are effective strategies to be used in cases with limited dataset.^38^ Transfer learning is a special case in which a CNN based DL model trained on one type of dataset or domain is re-purposed on another dataset. Transfer Learning demonstrates compelling results, particularly in cases where the data available for building the models is limited.^11, 38^ In the present study, 19 types of different CNN algorithms were trained, with each image in the training dataset augmented in 23 different ways. This increased the numbers of images in the training dataset and assured that the model was trained on various images, making it more robust and reliable to be used in clinical settings. The robustness was also evident because the prediction for external dataset images displayed better results in their original form than with augmentations. This indicates that in natural settings, apart from pixel changes, no query image augmentation is required.

We looked for any pattern between the DL predictions of ADoA and specific gene mutations. In all the mutation positive patients, ADoA could be detected in over 80% of images. This was in contrast to only 5% of the normal images deemed as having ADoA. Most gene mutation studies on animal models of glaucoma have provided insights into the pathogenesis of outflow channels in controlled experiments, on the other hand, in-vivo analyses of human eyes with rare disease-causing mutations provides a better understanding of the anatomical effects of these mutations. While the mutations in the *MYOC* gene are known to cause aggregation of the misfolded myocilin protein that, leads to TM cell toxicity and eventually death, there is no evidence in literature to suggest the role of the *MYOC* gene in the development of the angle. There is only one histopathological report of a JOAG patient with *MYOC* Tyr453His mutation, where no apparent changes of the TM or SC were noted though intense *MYOC* immune-reactivity was observed at the TM ^39^. Nevertheless, there is evidence to suggest that *MYOC* mutations are associated with goniodysgenesis. Cheng X^40^ et al. reported a 3 generation JOAG family with Pro370Leu mutation in the *MYOC* gene in all affected members who also had goniodysgenesis. This evidence is further strengthened by the reports of the association of *MYOC* gene mutations with congenital glaucoma.^17-19^ In contrast,*CYP1B1* related cases of PCG have been shown to have histopathological evidence of goniodysgenesis, involving not only the TM and SC but also the collector channels.^41^ In our study too, the subset of JOAG patients with *CYP1B1* gene mutations showed ADoA as predicted with DL models. We also found *FOXC1* and *LTBP2* mutations among our patients with no other features of either Axenfeld Reiger Syndrome (ARS) or zonular abnormalities classically associated with these gene mutations. Two cases of JOAG with *LTBP2* mutations have been described^42,43^, one by Saeedi et al. and the other, by our group. The *LTBP2* gene mutations are known to express a wide variety of ocular phenotypes (as with other monogenic disorders) ranging from primary trabecular meshwork dysgenesis to a Marfans like zonular disease. While *FOXC1* mutations have been commonly associated with ARS, they are also known to occur in adult onset POAG and JOAG.^44^ In our study, 2 unrelated patients harboured the same *FOXC1* frameshift mutation which is novel. Our findings demonstrate that probably different gene mutations affect different parts of the proximal outflow pathways. While *CYP1B1* and *LTBP2* was found to affect primarily the SC morphology, the *MYOC* and *FOXC1* were found to be associated with morphological variations in the TM, since the SC in the latter was normally developed.

The present study’s limitation is the fewer images in the training dataset (n=340) used for DL model building. However, we enhanced the input data by using augmentation techniques. Another limitation of the limited data set was our inability to correlate the gene mutations with the clinical severity of the disease, which was not within the ambit of our research since our study was focussed on evaluating the association gene mutations with DL predicted angle dysgenesis. Many images on ASOCT have to be discarded due to poor quality and image artefacts at the ICA area due to the reflectance from the superficial vessels. This would be taken care of, hopefully, in the newer generation machines, which would have better resolution too. Notwithstanding these limitations, the strength of the study lies in having addressed a crucial as well as unique issue of in vivo identification of angle dysgenesis using a very rare dataset of early onset POAG patients.

In conclusion, we have built a consensus based DL model to predict the presence or absence of ADoA. The validation on independent datasets and its correlation with the known gene mutations has highlighted the translational relevance of the model in clinical settings, as it could potentially be deployed in screening patients and their family members who could be picked up if they have angle dysgenesis.

## Data Availability

All data produced in the present study are available upon reasonable request to the authors

## Abbreviations and acronyms used

AI: Artificial Intelligence
ML: Machine Learning
DL: Deep learning
SC: Schlemm’s canal
SD-OCT: Spectral domain–optical coherence tomography
PCG: Primary congenital glaucoma
JOAG: Juvenile-onset open angle glaucoma
POAG: Adult-onset primary open angle glaucoma
TM: Trabecular meshwork
ICA: Iridiocorneal angle
IOP: Intraocular pressure
ASOCT: Anterior Segment Optical Coherence Tomography
ADoA: Angle Dysgenesis from the ASOCT
AUROC: Area under receiver operating characteristic curve

## Legends

Supplementary Fig 1: Flow chart of the analysis of anterior segment SD-OCT image

## Author contributions

VG, DG, SB conceptualized the study, analyzed and prepared the manuscript. SB developed the deep learning models. VG and BIS conducted the genetic studies and bio informatics. VG, SG, AS, KM and TV conducted patient recruitment and clinical studies. DG and MG reviewed and modified the final version.

