## Supplemental Table for "Deep learning based models to study the effect of glaucoma genes on angle dysgenesis in-vivo"

**Suppl Table 1: Architecture, optimized hyperparameters and performance of the three final classification models**

|  | **Model-1**  **(inceptionresnet-v2)** | **Model-2**  **(inceptionresnet-v2)** | **Model-3**  **(mobilenet-v2)** |
| --- | --- | --- | --- |
| **Input-images** | Iridocorneal angle (ICA) area | Trabecular meshwork (TM) area | Trabecular meshwork (TM) area |
| **Properties** | Layer:[824×1nnet.cnn.layer.Layer] Connections: [921×2 table] | Layer:[824×1nnet.cnn.layer.Layer] Connections: [921×2 table] | Layers: [154×1nnet.cnn.layer. Layer] Connections: [163×2 table] |
| **EPOCH** | 45 | 45 | 30 |
| **Learning_Rate** | 0.005 | 0.005 | 0.005 |
| **MiniBatchsize** | 64 | 64 | 32 |
| **Shuffle** | 'every-epoch' | 'every-epoch' | 'every-epoch' |
| **SolverName** | sgdm | sgdm | sgdm |
| **Internal testing dataset** | | | |
| **Total images** | 8160 | 8160 | 8160 |
| **Training dataset (98%)** | 7996 | 7996 | 7996 |
| **Testing dataset (2%)** | 164 | 164 | 164 |
| **Accuracy %** | 97.56 | 98.17 | 98.78 |
| **Sensitivity** | 0.964 | 0.97 | 0.987 |
| **Specificity** | 0.987 | 0.987 | 0.987 |
| **Precision** | 0.987 | 0.98 | 0.987 |
| **Recall** | 0.964 | 0.97 | 0.987 |
| **External independent validation dataset** | | | |
| **External dataset (n)** | 67 | 67 | 67 |
| **Accuracy %** | 83.58 | 74.6 | 80.6 |
| **Sensitivity** | 0.68 | 0.64 | 0.80 |
| **Specificity** | 0.92 | 0.78 | 0.80 |
