## Supplementary figures and images for "Deep learning based models to study the effect of glaucoma genes on angle dysgenesis in-vivo"

### Supplemental Fig 1

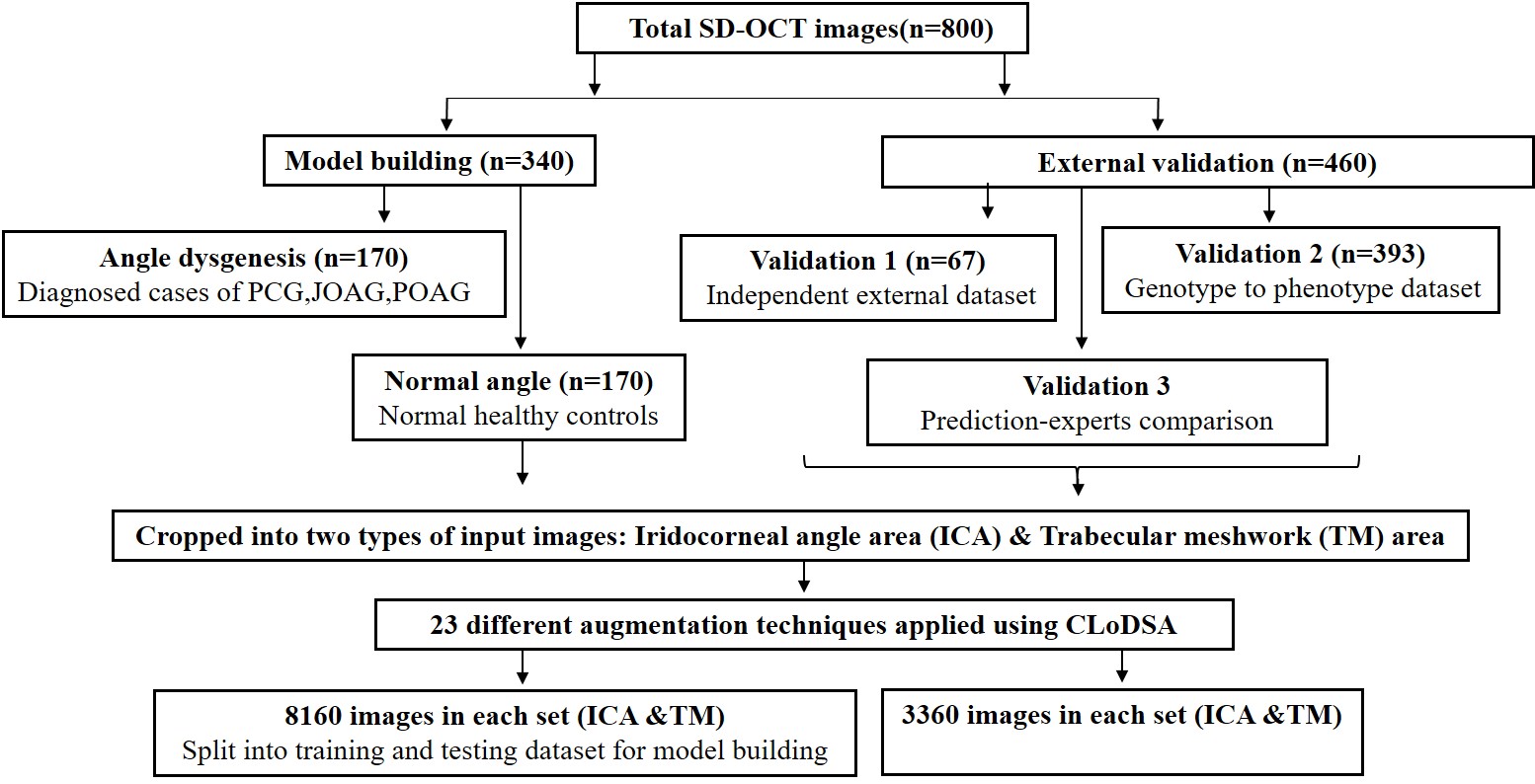
