## Supplemental information 1 for "Deep learning based models to study the effect of glaucoma genes on angle dysgenesis in-vivo"

*Material and Methods*

***Deep Learning***

*Augmentation technique*

To avoid overfitting, incorporate variability and increase the size of the datasets, 23 different types of augmentation techniques were performed on each image using the CloDSA tool, a freely available package.^12^ The different augmentation includes Horizontal flip, Vertical flip, Random Rotation, Rotation 45, Rotation 90, Rotation 120, Rotation 140, Rotation 160, Rotation 270, Average Blurring, Change HSV, Change LAB, Gamma correction, Gaussian blur, Raise blur, Raise green, Raise red, Raise saturation, Raise hue, Median blur, Salt and pepper, Sharpen and Translation. Finally, the total number of images after augmentations in each dataset increased to 8160, which were divided into training and testing sets for model building.

*Hardware and technologies used for AI models*

The entire model building experiments were carried out at a workstation with Intel (R) Xeon (R) W-2133 CPU @3.60 * 12 GZ processors, Graphics – Quadro P5000/PCle/SSE2, 64 GB RAM and CentOS- 7 (Core). The AI studies were performed using MATLAB 2021a, a high-performance language for technical computing developed by The MathWorks, Inc, USA. The four main MATLAB toolboxes, (image processing, computer vision, deep learning and parallel computing toolbox) were used.

***Genetic Evaluation***

*Exome sequencing*

Two ml blood drawn from the antecubital vein of patients were collected in EDTA (ethylene diamine tetra acetate) vial and DNA was isolated by salting out method. Exome capture was achieved using the Sure Select Human All Exon V5 (50.4 Mb) (Agilent Technologies, Santa Clara, California, USA) by following manufacturer’s instructions. Briefly, 1µg of genomic DNA was fragmented (150 –200 bp) and ligated to adapter primers, and then PCR amplified. Further, biotinylated RNA capture probes (~120 bp) were used for hybridization of amplified DNA-fragment libraries. Hybridised DNA/RNA was recovered by streptavidin-coated magnetic bead separation (Dynal, Invitrogen, Carlsbad, CA). Hybrid-capture libraries were amplified to add the sequencing primers and identifying tags and then subjected to paired-end (2 x 101 bp read length) multiplex sequencing. Captured DNA was eluted and then subject to flow-cell massively-parallel sequencing on a HiSeq2000System (Illumina, San Diego, CA). All samples were sequenced with a minimum of 100x coverage.

*Exome variant analysis*

All the Fast Q files were assessed for the quality of raw sequence data. Raw reads with poor base quality and sequence adapters were trimmed by trimmomatic. The clean reads with Phred score > 20 were mapped to the human reference genome build hg19/GRCh37 using Burrows-Wheeler Aligner (BWA). SAM tools was used for sorting of reads and duplicate reads was marked by Picard tools (http: //picard. sourceforge.net/). Genome analysis tool kit (GATK) v2.7.2) was used for calling variants. Further the variant calling format files (VCF) were annotated using Golden Helix VarSeq Software v.1.2.1 (Bozeman, MT). Target coverage and read-depth were reviewed by the Integrated Genomics Viewer (IGV, http://www. broadinstitute.org/igv/). Variant filtering was based on (a) read depth (DP) of >10 (b) genotype quality (GQ) score of >20 and (c) predicted missense or loss-of-function mutation.

Variants were prioritised by following steps. (1) Rare or novel variants (having minor allele frequency ≤1% ) including frame shift, splice, stop gain, stop loss or missense predicted to be damaging by at least two pathogenic prediction scores, were filtered. Minor allele frequencies (MAF) were accessed from public databases including 1000 Genome Project phase 3 ([www.1000genomes.org](http://www.1000genomes.org)),dbSNP Common 151 ([http://ncbi.nlm.nih.gov/SNP/), ExomeVariantServer](http://ncbi.nlm.nih.gov/SNP/),%20ExomeVariantServer) (EVS, http://evs.gs.washington.edu/EVS/) and (2) variants known to be associated with Glaucoma in published literature.
